# Evaluation of commercial anti-SARS-CoV-2 antibody assays and comparison of standardized titers in vaccinated healthcare workers

**DOI:** 10.1101/2021.08.24.21262475

**Authors:** Kahina Saker, Vanessa Escuret, Virginie Pitiot, Amélie Massardier-Pilonchéry, Stéphane Paul, Bouchra Mokdad, Carole Langlois-Jacques, Muriel Rabilloud, David Goncalves, Nicole Fabien, Nicolas Guibert, Jean-Baptiste Fassier, Antonin Bal, Sophie Trouillet-Assant, Mary-Anne Trabaud

## Abstract

With the availability of vaccines, commercial assays detecting anti-SARS-CoV-2 antibodies (Ab) evolved towards quantitative assays directed to the spike glycoprotein or its receptor binding domain (RBD). The main objective of the present study was to compare the Ab titers obtained with quantitative commercial binding Ab assays, after 1 dose (convalescent individuals) or 2 doses (naïve individuals) of vaccine, in healthcare workers (HCW).

Antibody titers were measured in 255 sera (from 150 HCW) with 5 quantitative immunoassays (Abbott RBD IgG II quant, bioMérieux RBD IgG, DiaSorin Trimeric spike IgG, Siemens Healthineers RBD IgG, Wantai RBD IgG). One qualitative total antibody anti RBD detection assay (Wantai) was used to detect previous infection before vaccination. The results are presented in binding Ab units (BAU)/mL after application, when possible, of a conversion factor provided by the manufacturers and established from a World Health Organization (WHO) internal standard.

There was a 100% seroconversion with all assays evaluated after two doses of vaccine. With assays allowing BAU/ml correction, Ab titers were correlated (Pearson correlation coefficient, ρ, range: 0.85-0.94). The titer differences varied by a mean of 10.6% between Siemens and bioMérieux assays to 60.9% between Abbott and DiaSorin assays. These results underline the importance of BAU conversion for the comparison of Ab titer obtained with the different quantitative assays. However, significant differences persist, notably, between kits detecting Ab against the different antigens.

A true standardization of the assays would be to include the International Standard in the calibration of each assays to express the results in IU/mL.

## Introduction

Since the end of 2020, severe acute respiratory syndrome coronavirus-2 (SARS-CoV-2) vaccines have become available worldwide with the aim of achieving herd immunity to control the pandemic. Vaccine immunity involves both cellular and humoral pathways. Cellular immunity is not easy to assess on a large scale, as is the neutralizing humoral response owing to requirement for a biosafety level 3 (BSL3) containment laboratory. The evaluation of vaccine effectiveness therefore mainly relies on high throughput serological tests to assess individual humoral immunity as well as monitoring SARS-CoV-2 seroprevalence (1).

To effectively utilize measurements of binding antibodies (Ab) as indicators of vaccine effectiveness, several conditions must be met. First, binding Ab assays should be quantitative; second, titers should be consistent between different assays; third, binding Ab titers should correlate with neutralizing Ab titers; fourth, the minimum binding Ab titer associated with virus neutralization must be found; and fifth, the association between neutralizing Ab and vaccine protection must be demonstrated. It can be considered that the first and fifth conditions have been met given that commercial tests for the quantitative detection of binding Ab have been developed (2–8), and that the role of neutralizing Ab in the infection protection have been demonstrated in animals and humans (9-12). This is not the case for the other conditions; in particular, the second point is of importance for widespread evaluation of vaccines, but until now, Ab titers were often expressed as an index or unit with regard to an internal standard that differs between manufacturers. Recently, the World Health Organization (WHO) has developed an international standard (13) against which each supplier can standardize their assay, allowing comparability of titers between kits. The present study was conducted to evaluate the performance of commercial antibody assays in detecting vaccination-associated anti-SARS-CoV-2 Ab seroconversion; the main objective was to compare Ab titers from quantitative assays after conversion of titers with the conversion factor obtained using the WHO standard and provided by each manufacturer.

### Materials and Methods Antibody binding assays

Six CE-marked Ab binding assays, validated by each manufacturer, were investigated according to the protocol recommended by each manufacturer (the characteristics of the assays are summarized in Table 1). Five were quantitative: Siemens Healthineers (Erlangen, Germany) Atellica® IM SARS-CoV-2 IgG (sCOVG; used in routine in our laboratory), DiaSorin (Saluggia, Italy) Liaison® SARS-CoV-2 TrimericS IgG, bioMérieux (Marcy l’Etoile, France) Vidas® SARS-CoV-2 IgG (clinically used for confirmation if necessary), Abbott (Abbott park, Il, USA) Architect SARS-CoV2 IgG II Quant, and Wantai (Beijing, China) SARS-CoV-2 IgG assays. The Wantai SARS-CoV-2 total antibody assay is qualitative and was selected to detect a previous infection before vaccination based on its better sensitivity in infected individuals compared to other commercial qualitative assays we have evaluated in a previous study (14). The First International Standard developed by the WHO (National Institute for Biological Standards and Control code: 20/136) corresponds to lyophilized pooled plasma from patients who had been infected with SARS-CoV-2; after reconstitution, the solution contains 1000 binding antibody units (BAU) per mL (13).

**TABLE 1:**
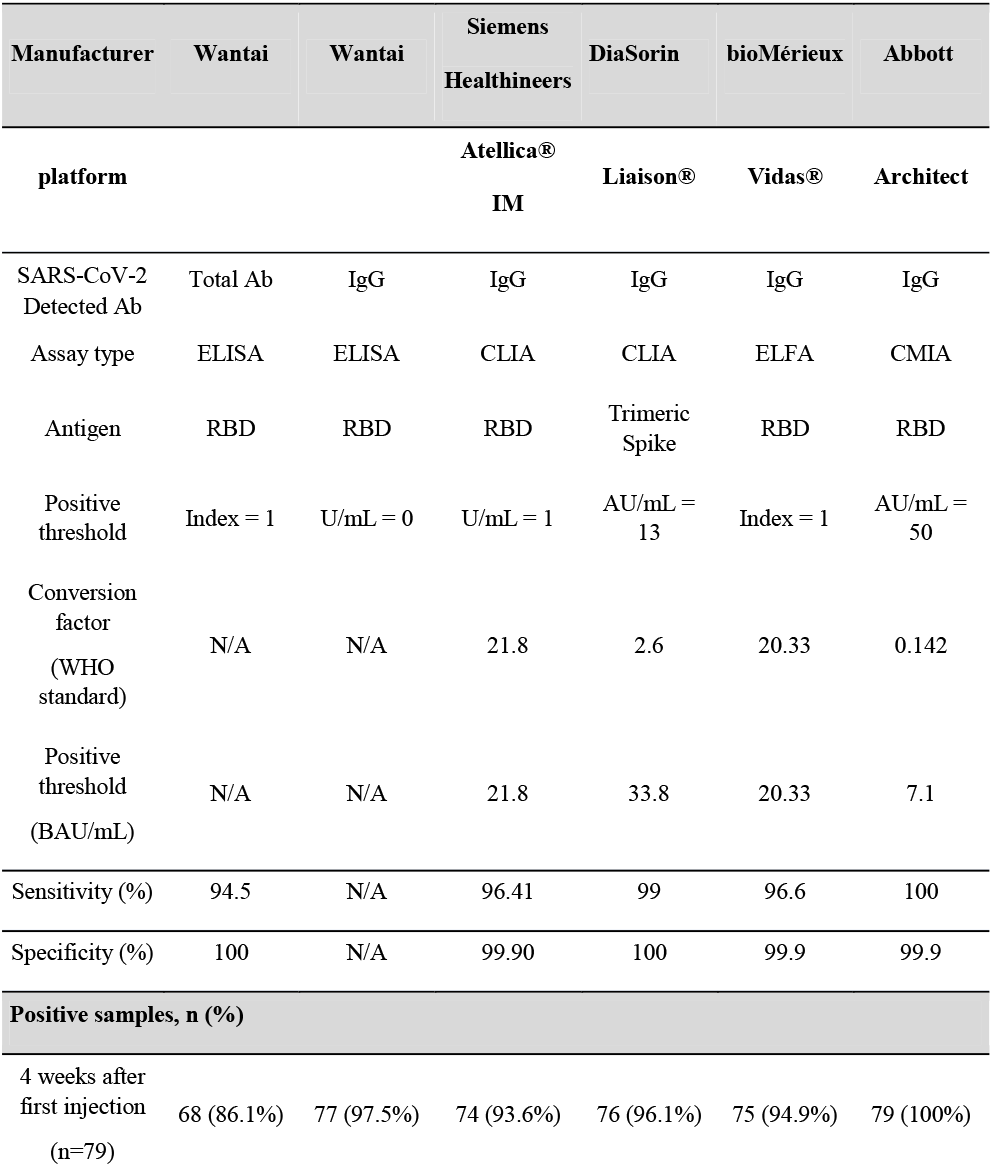
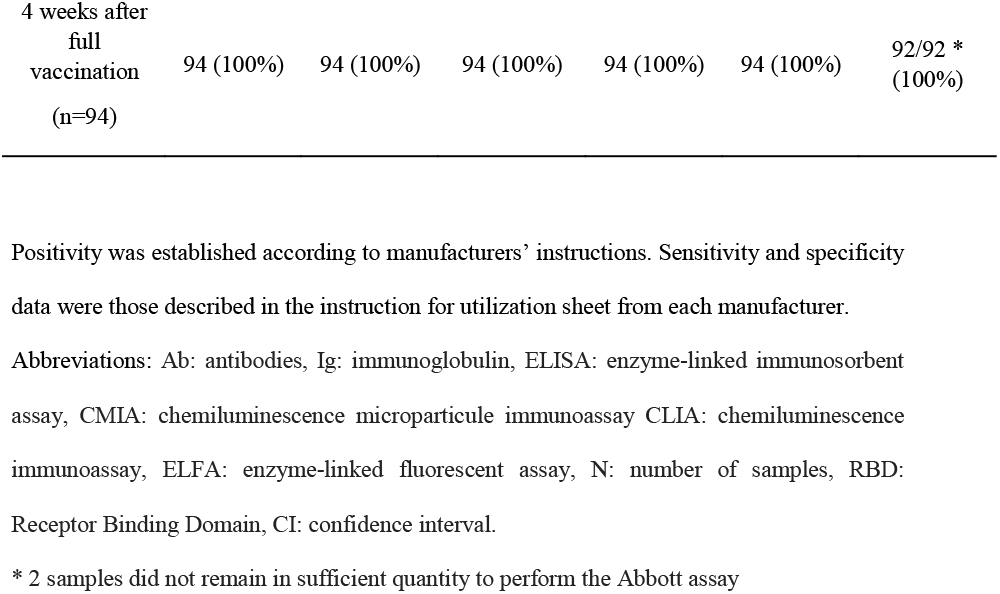
Performance of six commercial anti SARS-CoV-2 antibody assays.

For conversion of titers obtained using the quantitative assays, the concentrations expressed in arbitrary units per mL, or index according to the assay (Table 1), were converted to BAU/mL using the conversion factors provided a posteriori (not included in the main procedure but as a separate document – either by electronic or postal mail) by the manufacturer (with the exception of the Wantai SARS-CoV-2 IgG assay for which the conversion factor was not available and presented here only to compare the positivity rate between assays); these were 21.8 for the Siemens assay, 2.6 for the DiaSorin assay, 20.33 for the bioMérieux assay, and 0.142 for the Abbott assay (considering that 1 BAU/mL = conversion factor x AU/mL or index). Samples with results above the upper limit of quantification were tested again after dilution (1/5 when above 3270 BAU/mL for the Siemens assay, 1/20 when above 2080 BAU/mL for the DiaSorin assay, 1/20 when above 18 index for the bioMérieux assay, and 1/2 when above 5680 BAU/mL for the Abbott assay).

### Samples from the study population

A prospective longitudinal cohort study was conducted at the laboratory associated with the national reference center for respiratory viruses (University Hospital of Lyon, France). Healthcare workers, excluding pregnant women (HCW; n=150) who were scheduled to receive 2 doses of Pfizer BioNtech vaccine (n=94; BNT162b2/BNT162b2; 78% female; median age 48.5 [range: 21-76] years) or 1 dose of AstraZeneca vaccine followed by 1 dose of Pfizer BioNtech vaccine (n=56; ChAdOx1/BNT162b2; 70% female; median age 33.5 [range: 21-55] years) were included. Blood samples were collected i) before the first dose of vaccine, ii) before the second injection of vaccine corresponding to 4 weeks after the first dose for participants vaccinated with 2 doses of Pfizer BioNtech vaccine (median delay of 28 [range: 21-37] days) or 12 weeks for those vaccinated with the AstraZeneca vaccine (median delay of 85 [range: 84-97] days), and iii) 4 weeks after the full vaccination. The pre-vaccination blood sample was used to document a previous SARS-CoV-2 infection. Among the participants, 26 who were previously infected with SARS-CoV-2 (convalescent group; 17.4%) had only one vaccine injection (Pfizer BioNtech, n=15 or Astra Zeneca, n=11); for these the second sample was omitted. Three participants were infected with SARS-CoV-2 between the 2 doses. Serum samples were prepared from 5 mL of whole blood collected in BD Vacutainer^®^ Serum Separator Tubes II Advance (Beckon Dickinson Diagnostics). After collection, tubes were shaken gently and serum allowed to clot for a minimum 30 min at room temperature to obtain total coagulation, followed by centrifugation at 2,000 g for 10 min. Serum removed from gel was stored at -80°C until serological assays were performed.

Written informed consent was obtained from all participants; ethics approval was obtained from the regional review board for biomedical research in April 2020 (*Comité de Protection des Personnes Sud Méditerranée I*, Marseille, France; ID RCB 2020-A00932-37), and the study was registered on ClinicalTrials.gov (NCT04341142).

### Statistical analyses

Results were expressed by the median and interquartile range [IQR]. Paired comparison between assays was performed using the Wilcoxon test. The correlation between concentrations obtained by each assay was investigated using Pearson correlation coefficients and 95% confidence interval (CI). To estimate proportional bias between two methods, Passing and Bablok regression was used and the regression line equation was calculated from the two data sets. The Bland-Altman method was used to measure the mean difference and 95% limit of agreement between log-transformed concentrations obtained with each assay. Statistical analyses were conducted using GraphPad Prism® software (version 8; GraphPad software, La Jolla, CA, USA). A p-value <0.05 was considered statistically significant.

## Results

In the first part of the study, the performances of the six assays were compared to verify whether the ability to detect anti-SARS-CoV-2 antibodies of the Wantai total Ab assay, previously found as the most sensitive post-infection compared to other commercial qualitative assays (14), was similar after vaccination, and whether the sensitivity of qualitative and quantitative assays were also similar. The sera collected from patients scheduled to receive only Pfizer BioNtech vaccine (two doses 3 to 4 weeks apart, n=79) were used for this evaluation. Four weeks after the first injection, the proportion of positive samples was over 90% for all assays except the Wantai assay detecting total antibodies (86.1%). Only one sample was negative with all assays. Six samples were negative with only one assay (5 with the Wantai total Ig assay, 1 with the Wantai IgG assay). Four samples were negative with 3 to 5 assays. In all of these cases the signals from positive assays were low. Four weeks after the second injection anti SARS CoV-2 antibodies were detected for all participants with all the assays (Table 1).

The second part of this study was to compare Ab titers after BAU/mL conversion; for this, only the assays adapted or developed for the quantification of Ab were tested. The Siemens, DiaSorin, bioMérieux and Abbott assays were compared using sera samples collected before the second injection of vaccine and those collected after full vaccination, and for which assays gave a positive quantitative result (255 samples). The median [IQR] values obtained were 744.9 [108.1; 2482] BAU/mL for the Siemens, 1240 [262.6; 3370] BAU/mL for the DiaSorin, 951.4 [142.5; 2314] BAU/mL for the bioMérieux, and 768.9 [100.8; 1916] BAU/mL for the Abbott assays; there was a significant difference in median titers between DiaSorin and Siemens (p<0.0001), between DiaSorin and bioMérieux (p<0.0001), as well as between Abbott and each assay (p<0.0001). The difference in median titers between Siemens and bioMérieux assays was not significant (Figure 1). There was a strong correlation between assays (Table 2).

**FIG 1:**
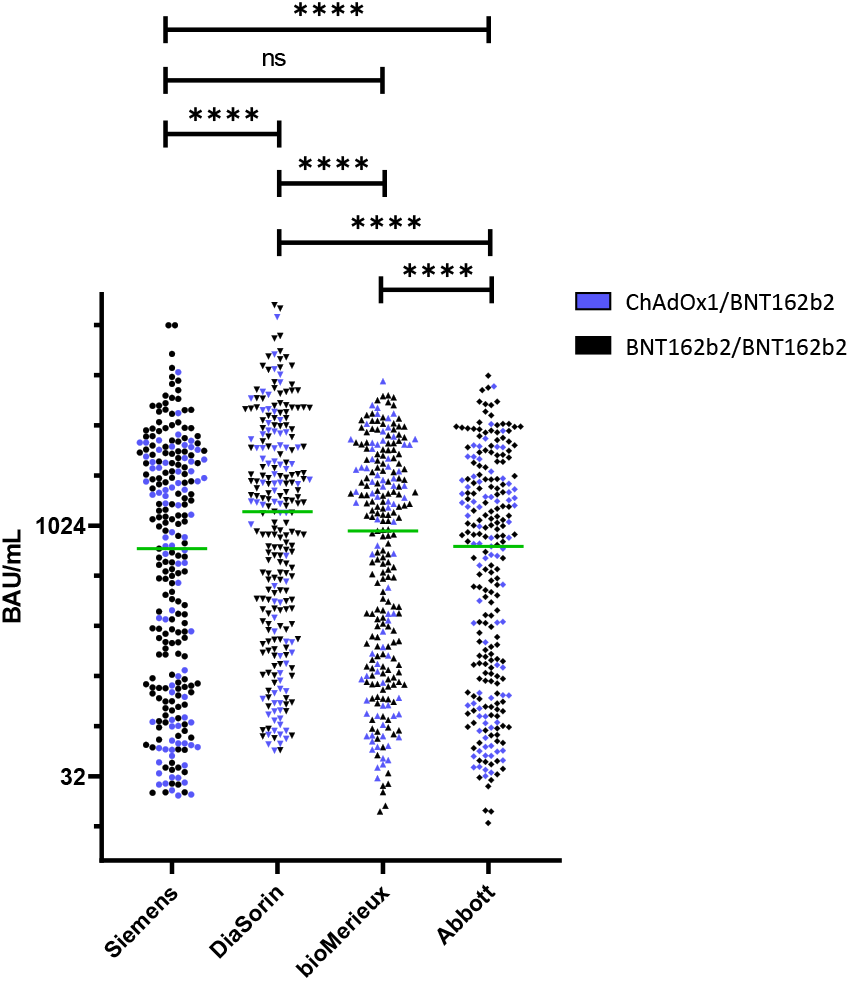
Comparison of anti SARS-CoV-2 antibodies concentration (BAU/mL) between all assays in sera collected from vaccinated subjects. The statistical difference was evaluated by Wilcoxon’s test. Comparison of median titers between Siemens, DiaSorin, bioMérieux, and Abbott assays. BAU/mL: Binding Antibodies Unit/mL. ****p<0.0001. Data from patients scheduled to be vaccinated with 2 doses of Pfizer BioNtech vaccine (black) or with 1 dose of AstraZeneca vaccine followed by 1 dose of Pfizer BioNtech (blue) are presented.

**TABLE 2:**
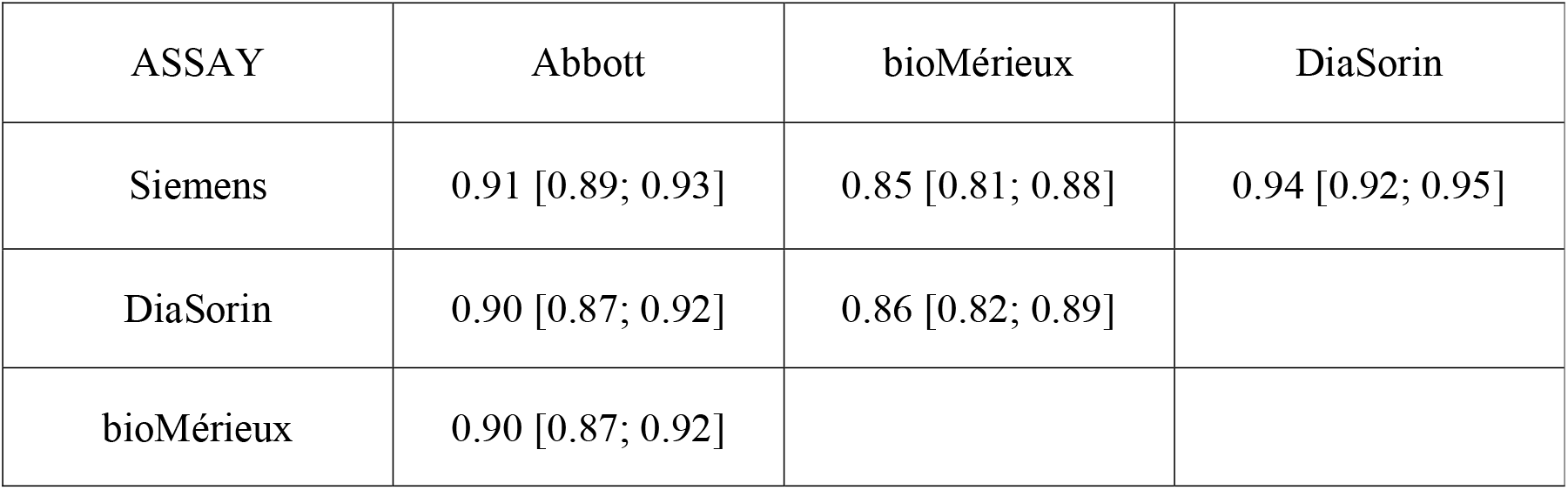
Pearson correlation coefficient (ρ [IQR]) between each assay.

| ASSAY | Abbott | bioMérieux | DiaSorin |
| --- | --- | --- | --- |
| Siemens | 0.91 [0.89; 0.93] | 0.85 [0.81; 0.88] | 0.94 [0.92; 0.95] |
| DiaSorin | 0.90 [0.87; 0.92] | 0.86 [0.82; 0.89] |  |
| bioMérieux | 0.90 [0.87; 0.92] |  |  |

The slopes of the Passing and Bablock regression curves were calculated by pairwise comparison of each assay. The deviation from the perfect correlation was significantly greater between DiaSorin and each of the other assays than between each of these other assays. In most cases, there was a proportional difference between assays (Figure 2). In particular, DiaSorin assay values reached higher levels than the other methods; 11 samples had a value above 10,000 BAU/ml while only three reached this threshold with the Siemens assay and none with the BioMérieux and Abbott assays.

**FIG 2:**
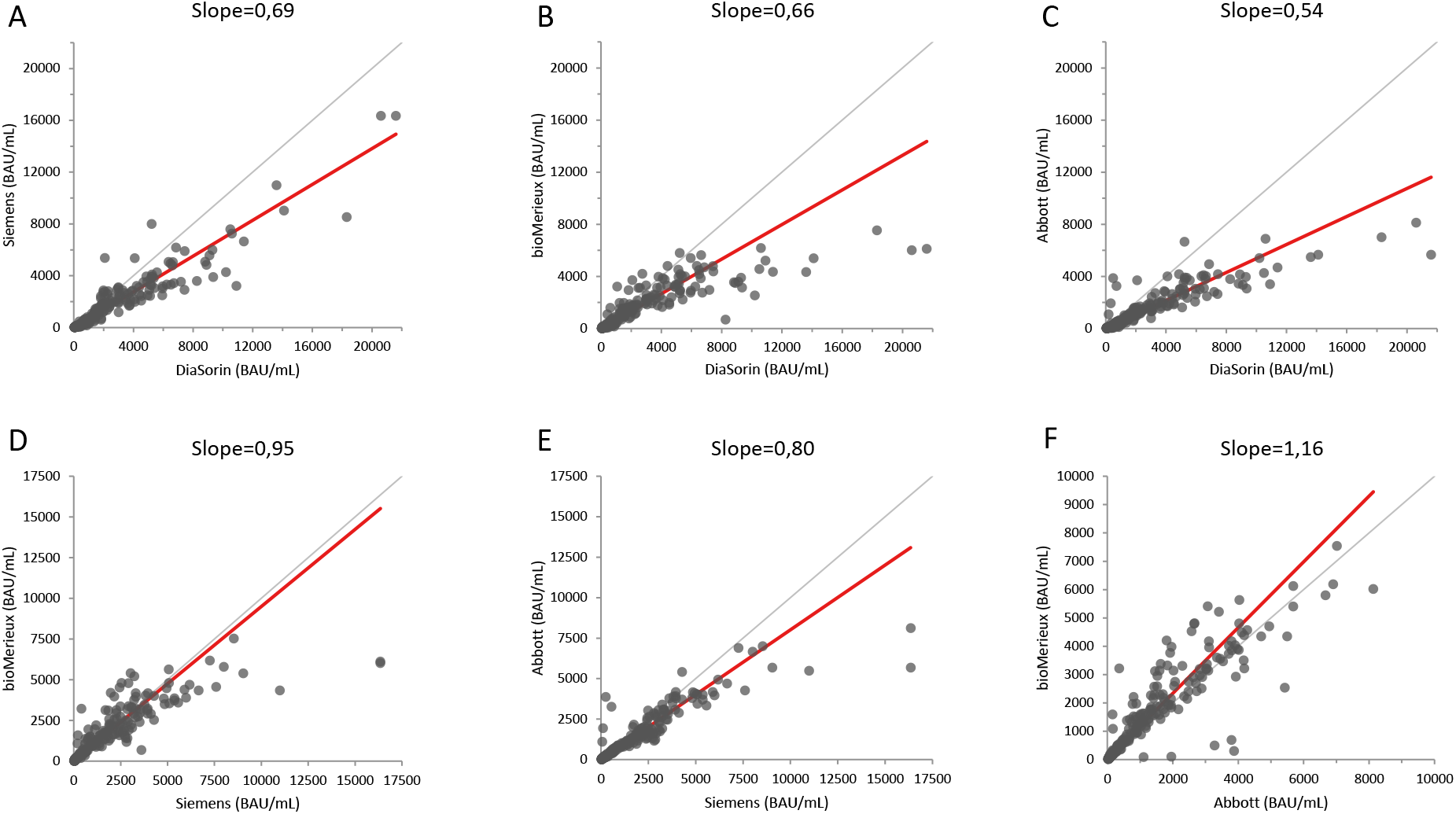
Passing and Bablok regression analyses using the Siemens, DiaSorin, bioMérieux, and Abbott assays. A, The Siemens assay compared to the DiaSorin assay. B, bioMérieux compared to DiaSorin. C, Abbott compared to DiaSorin. D, Siemens compared to bioMérieux. E, Siemens compared to Abbott. F, Abbott compared to bioMérieux.

According to the Bland-Altman method there was a mean [95%CI] difference in titers expressed as BAU concentrations of 50.9% [46.4%; 55.3%] between the Siemens and DiaSorin assays, 40.4% [35.5%; 45.3%] between bioMérieux and DiaSorin assays, 60.9% [55.9%; 66%] between Abbott and DiaSorin assays, 10.6% [5.8%; 15.4%] between Siemens and bioMérieux assays, 12.1% [16.2%; 8%] between Siemens and Abbott assays, and 22.3% [17.5%; 27.1%] between Abbott and bioMérieux assays (Figure 3).

**FIG 3:**
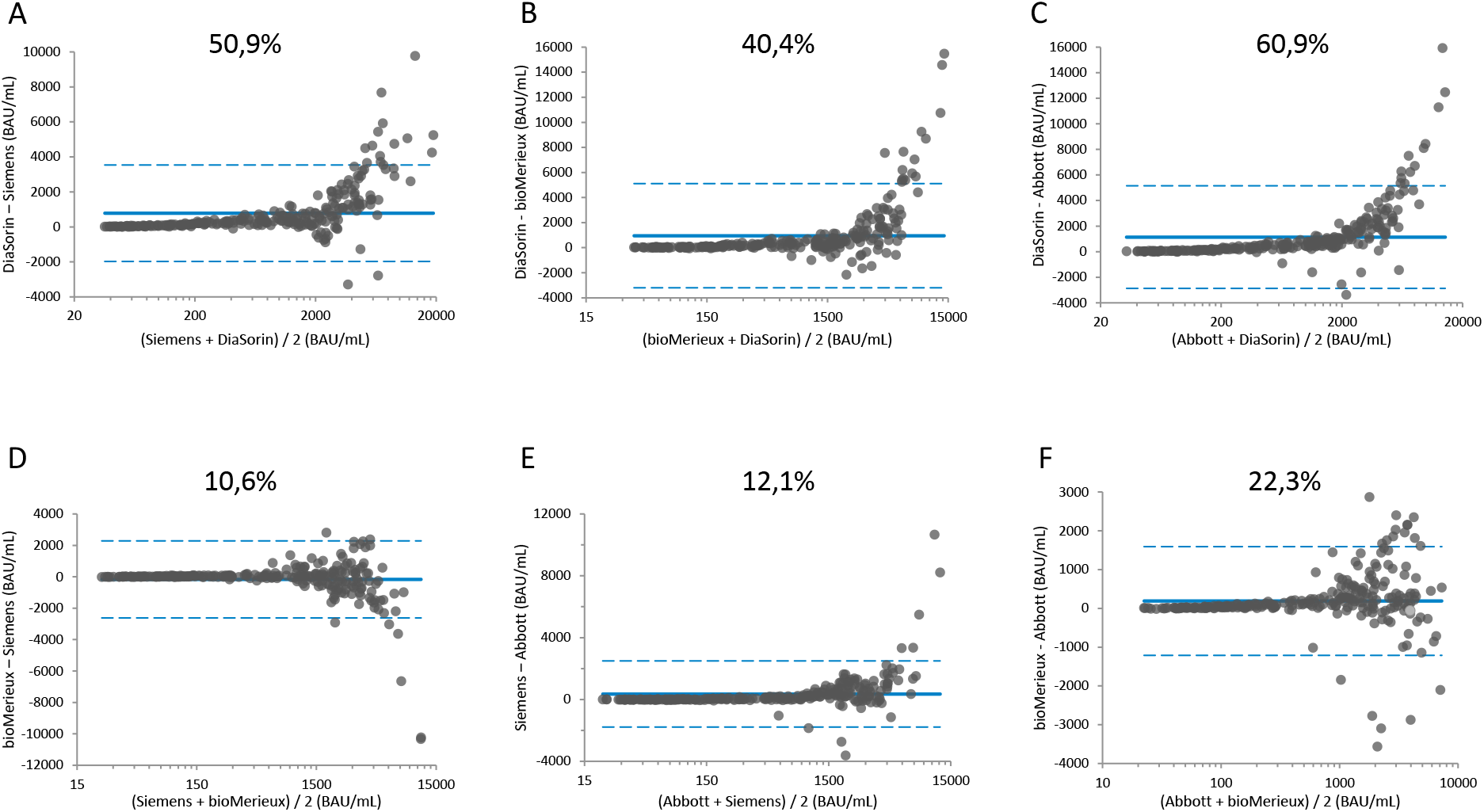
Bland-Altman plots comparing agreement between concentrations determined using the Siemens, DiaSorin, bioMérieux, and Abbott assays. A, The Siemens assay compared to the DiaSorin assay. B, bioMérieux compared to DiaSorin. C, Abbott compared to DiaSorin. D, Siemens compared to bioMérieux. E, Siemens compared to Abbott. F, Abbott compared to bioMérieux. The solid blue line represents the bias between assays, the dashed blue lines represent 95%CI.

## Discussion

In the present cohort of vaccinated HCW, the performance of qualitative serological assays, developed for diagnostic purposes at the beginning of the pandemic, as well as those adapted or developed when the vaccine became available, were similar. However, assays developed for detecting a past infection are not useful to monitor vaccination effectiveness since they are not quantitative or could not be compared to others. The more important finding of the present study was that the quantitative assays, whose results could be standardized to BAU/ml, produce results that are correlated to each other. The Ab titers obtained with the DiaSorin assay, although correlated with those of the other assays, remained higher after conversion using the WHO standard. This may be related to difference in the antigens targeted, as the DiaSorin assay is the only one evaluated in this study, to use the trimeric spike protein (2). The WHO standard was obtained from the plasma of convalescent patients (13) and must therefore contain antibodies against numerous epitopes, thus the DiaSorin assay is probably capable of reacting with more antibodies than assays detecting only antibodies specific for RBD.

There are currently very few reports that have examined Ab binding assays with the use of the WHO standard (3, 7). Perkman et al. (7) compared four assays detecting binding Ab, including the Liaison anti SARS-CoV-2 trimeric S IgG from DiaSorin and the Architect anti SARS-CoV-2 RBD IgG from Abbott, used in the present study. In vaccinated individuals, after the first dose of vaccine, titers varied significantly between the assays, but they indicated that the recalculation in BAU/ml with the conversion factor given by the manufacturer did not solve error problems between tests. However, the assays compared were more different in their format than those investigated herein: total anti-RBD Ig versus anti-RBD IgG, anti-monomeric spike IgG or anti-trimeric spike IgG. Interestingly, comparison of Abbott and DiaSorin assays found, as was the case herein, higher titers for the DiaSorin assay. Later, Bradley et al. (3) performed linear regressions from sample dilutions of the WHO standard to determine a detection limit in international units (IU) per ml for each test; the authors confirmed the linearity of the Abbott anti-SARS-CoV-2 IgG II Quant assay over the analytical measurement interval but the conversion factor found seemed to be higher than indicated by the manufacturer (1 IU/ml gave 6,1 arbitrary units/ml while Abbott indicated that 1 IU correspond to 7.1 arbitrary units). Taken together, these data suggest that there are remaining differences between assays after conversion in BAU/ml and that this could be due to the incorrect adjustment of the correction factor by the manufacturers. The next step for a true harmonization would therefore be to use the international standard to calibrate each assay instead of applying a conversion factor to a result obtained with an assay previously calibrated with an internal standard (15).

A limitation of this study is the absence of specificity analysis. However, assay specificity analyses have been performed by manufacturers and independent groups (2, 4–6) showing specificity ≥99% for all the quantitative assays. A point that may be also considered as a limitation is the choice to use the conversion factors obtained by the manufacturers without testing the WHO standard ourselves; but the aim of the present study was to evaluate these assays under the conditions offered by the manufacturers to all their customers. In addition, not all commercial quantitative anti-SARS-CoV-2 Ab assays were evaluated, limiting the scope of the conclusions. Furthermore, neutralizing Ab were not investigated that could have helped determine whether anti-RBD or anti-spike assays are the most correlated with virus neutralization. However, before investigating this, harmonization of neutralizing Ab titers is also necessary to determine a common threshold from which vaccine protection could be predicted, allowing then to find the corresponding threshold with high throughput binding Ab assays. A study comparing different cell-based assays (with either live or pseudotyped viruses) to measure neutralization in vitro is rather reassuring, although differences were found according to the viruses used for pseudotyping (16). However, comparison of cell-based assays with surrogate virus neutralization tests (sVNT) that are based on ELISA, and measuring the competition of Ab and RBD for the binding to ACE, the cellular entry receptor of the virus, did not find good agreement; this is inconvenient, as these assays could be promising given that they have potential for large-scale.

In conclusion, the evaluated assays correlated well with each other but a difference in titers remained after adjustment to the same International Standard. Thus, the titer harmonization is not yet completely achieved, but it is better between assays detecting the same Ab against the same antigen than between assays with different targets.

## Data Availability

The data that support the findings of this study are available from the corresponding author, STA, upon reasonable request.

## Acknowledgements

This study was supported by Hospices Civils de Lyon and Fondation des Hospices Civils de Lyon. The respective suppliers kindly provided all the serological kits used in this study.

We thank all the staff members of the occupational health and medicine department of the Hospices Civils de Lyon who contributed to the sample collection. We thank all Clinical Research Associates, for their excellent work. We thank Karima Brahima and all the members of the clinical research and innovation department for their reactivity (DRCI, Hospices Civils de Lyon). Human biological samples and associated data were obtained from NeuroBioTec (CRB HCL, Lyon France, Biobank BB-0033-00046). We thank all the technicians from the virology laboratory of the Hospices Civils de Lyon who performed the assays on the automated platforms. Lastly, we thank all the healthcare workers for their participation in this clinical study, and Philip Robinson for critical reading of the manuscript and language revision.

